# Age-related differences in cancer relative survival in the US: a SEER-18 analysis

**DOI:** 10.1101/2022.09.02.22279479

**Authors:** Diana R Withrow, Brian D Nicholson, Eva JA Morris, Melisa L Wong, Sophie Pilleron

## Abstract

Cancer survival has improved since the 1990s, but to different extents across age groups, with a disadvantage for older adults. We aimed to quantify age-related differences in relative survival (RS - one-year, and one-year conditioning on surviving one year) for 10 common cancer types by stage at diagnosis. We used data from 18 United States Surveillance Epidemiology and End Results cancer registries and included cancers diagnosed between 2012-2016 followed until December 31, 2017. We estimated absolute differences in RS between the 50-64 age group and the 75-84 age group. The smallest differences were observed for prostate and breast cancers (1.8%-points [95% confidence interval (CI):1.5-2.1] and 1.9%-points [95%CI:1.5-2.3], respectively). The largest was for ovarian cancer (27%-points, 95%CI:24-29). For other cancers, differences ranged between 7 (95%CI:5-9, esophagus) and 18%-points (95%CI: 17-19, pancreas). Except for pancreatic cancer, cancer type and stage combinations with very high (>95%) or very low (<40%) 1-year RS tended to have smaller age-related differences in survival than those with mid-range prognoses. Age-related differences in one-year survival conditioning on having survived one-year were small for most cancer and stage combinations. The broad variation in survival differences by age across cancer types and stages, especially in the first year, age-related differences in survival are likely influenced by amenability to treatment. Future work to measure the extent of age-related differences that are avoidable, and identify how to narrow the survival gap, may have most benefit by prioritizing cancers with relatively large age-related differences in survival (e.g., stomach, esophagus, liver and pancreas).

**Novelty and Impact:** In this analysis of United States population-based cancer registry data, age-related differences in cancer survival varied widely, ranging from less than 1% absolute difference in localized breast and prostate cancer survival to over 30% absolute difference in localized pancreatic cancer survival. Focused efforts to reduce age-related differences in cancer survival may have greatest impact by prioritizing cancer site and stage combinations with the widest differences.

## Introduction

In the United States (US) in 2017, nearly half a million adults aged 75 and over were diagnosed with cancer,^1^ and around 260,000 cancer deaths occurred in the same age group.^2^ As the US population ages, more cancer cases and deaths will occur in older individuals.^3^ The International Cancer Benchmarking Partnership (ICBP) study in seven high-income countries with similar health care systems (excluding the US) recently showed that cancer survival has improved since the 1990s, but not similarly across age groups and cancer types. Even after taking into consideration differences in life expectancy, older adults have experienced smaller improvements in survival than younger people resulting in widening differences in cancer survival across age groups.^4,5^ Outside ICBP countries, the magnitude and pattern of age-related differences in cancer survival are poorly documented.

Up-to-date information about the pattern of age-related gaps in cancer survival is required to motivate and direct efforts to understand and, hence, improve cancer outcomes for older adults. Stage is an important predictor of survival, but among those studies that have measured age-related differences in cancer survival,^5-11^ those that have stratified by stage tend to be focused on a single cancer type.^8-11^ Comparing patterns in age-related differences in relative survival by stage across multiple cancer types with different prognoses, however, can provide insight and generate hypotheses as to how age-related differences in survival arise.

In this study, we describe age-related differences in one-year relative survival (RS) and one-year RS conditioning on surviving one year from diagnosis by stage across 10 cancer sites, namely, prostate, breast, rectum, colon, ovary, esophagus, stomach, liver, lung, and pancreas in adults aged over 50 years. RS is a measure of net survival, that is a cancer survival in the absence of other causes of death. Together, these ten cancer sites accounted for nearly 60 percent of all cancers diagnosed in US adults aged 50 and older in 2016.

## Methods

### Data Source

We included all first primary prostate, breast, rectum, colon, ovary, esophagus, stomach, liver, lung, and pancreas cancers diagnosed between 2012 and 2016 from 18 US Surveillance Epidemiology and End Results (SEER) cancer registries and followed for vital status through December 2017. The 18 SEER registries are located in California (San Francisco/Oakland, San Jose/Monterey, Los Angeles, Greater California), Connecticut, the Detroit metropolitan area, Hawaii, Iowa, New Mexico, Seattle, Utah, Georgia (Atlanta, rural Georgia and Greater Georgia), Alaska (restricted to Alaska Natives), Kentucky, Louisiana, and New Jersey.^12,13^ Together, these population-based registries cover nearly 30% of the US population.^12^

Seven of the included cancer sites were chosen to be comparable with the ICBP study results.^4^ Prostate, breast and liver cancers were added due to their high contribution to cancer incidence and mortality. Because of their high survival and use of surgery as a primary treatment modality, breast and prostate cancers also provide an interesting contrast to the other cancers included from which to generate hypotheses about drivers of age differences.

Cancers were originally coded using ICD-O-3 and grouped according to the SEER ICD-O-3/WHO 2008 definitions.^14^ As our primary interest was in age-related differences in survival, we included only cases diagnosed in patients aged over 50. We excluded cancers that were registered based on death certificate or autopsy only (1.5%), were non-malignant (6.4%), were missing age (0.04%), or with disagreement between vital status and survival time (0.4%).

### Statistical analysis

We estimated 1-year RS and 1-year RS conditioning on surviving 1 year from diagnosis using the Ederer II method. Relative survival is a metric of net survival, which estimates the probability of survival from cancer in the absence of other causes of death. It is especially useful for comparing survival between groups for whom background mortality differs since it is estimated based on the ratio of the observed survival to the expected survival of individuals of similar demographics in the general population.^15,16^

Expected mortality was estimated from life tables stratified by county, age, year, sex, race/ethnicity and county-level socioeconomic status (SES) index.^17^ The SES index is a composite variable based on seven county-level SES attributes collected by the American Community Survey.^18^

RS estimates were stratified by age (50-64, 65-74, 75-84, and 85-99 years), and stage (SEER Summary Stage, categorized as localized, regional, distant, and unknown/unstaged).^19^ We excluded patients aged 100+ as expected life tables extend only to age 99.^20^ Age-related differences in 1-year RS were assessed using the absolute differences in RS between the 50 to 64 age group and the 75 to 84 age group. We estimated 95% confidence interval around differences of RS using Monte Carlo simulations assuming the complementary log-log transformed RS values to be normally distributed.^21^ For each difference considered, the confidence limits were obtained by taking the 2.5 and 97.5 percentiles of the empirical RS difference distribution resulting from 100,000 random draws in the complementary log-log transformed RS values distribution.

If improvements in cancer survival are to be made, they are more likely to be observed at a population level among patients aged 75 to 84 compared to those 85 to 99. Therefore, the 75 to 84 age group was used as reference to calculate absolute differences in survival.

SEER data are publicly available and therefore ethics approval was not required. Data and relative survival estimates were retrieved through SEER*Stat software version 8.3.9 (National Cancer Institute). Data analysis was conducted between June and August 2021 by DW and SP. The estimation of confidence intervals around differences and plotting of results were performed using R statistical software (version 4.0.2; R Development Core Team, 2020).

## Results

We included a total of 844 296 people diagnosed with cancer between the ages of 50 to 99 between 2012 and 2016. Figure 1 presents the distribution of stage at diagnosis by age groups for the ten cancer sites. Over half of prostate and breast cancers and about half of liver cancers were diagnosed at localized stage while less than a quarter of ovarian, esophageal, lung and pancreatic cancers were diagnosed early. The proportion of cancers of unknown stage increased after the age of 75 and was largest in the group aged 85 to 99, with percentages ranging from 6% for breast cancer to 29% for esophageal cancer in this age group.

**Figure 1.**
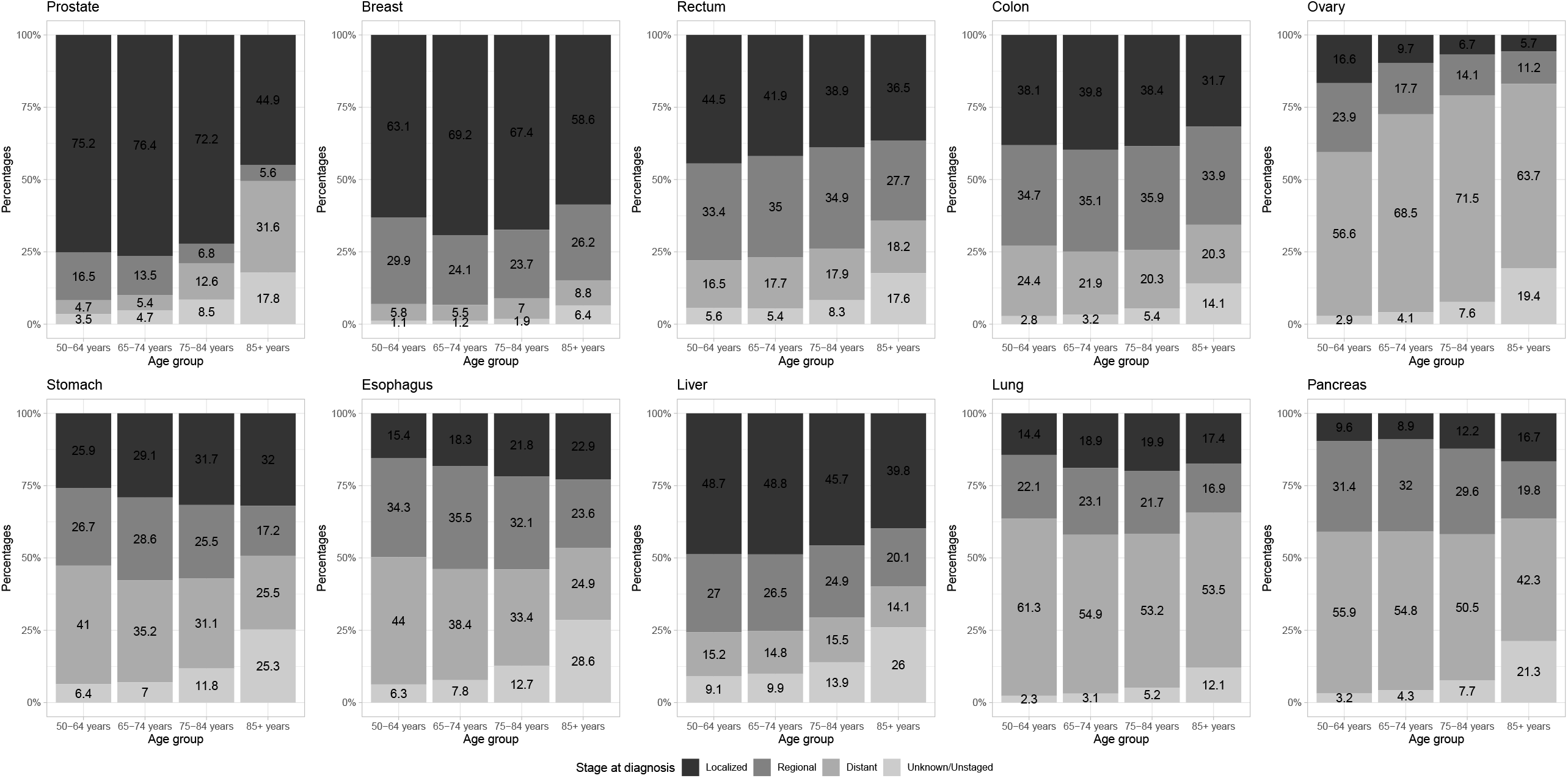
Distribution of stage at diagnosis by age groups and cancer types.

### Overall

One-year RS estimates were highest for prostate and breast cancers (>80% across age groups) and lowest for pancreatic cancer (43% in 50 to 64 age group and 25% in 75 to 84 age group, Table 1). Prostate, breast, colon, rectal and ovarian cancers all had relatively good prognosis in the youngest age group (1-year RS ≥86% in persons aged 50 to 64 years) whereas stomach, esophageal, liver, lung, and pancreatic cancers had relatively poor prognosis (42% to 63% in persons aged 50 to 64 years). For localized and regional prostate and breast cancers, 1-year RS remained high (>95%) until age 85, whereas survival for other cancer sites tended to decrease with increasing age. For unknown/unstaged cancers, 1-year RS fell between the RS observed for regional and distant stages, except for ovarian cancer for which unknown cancers had poorer RS than distant cancers.

**Table 1.**
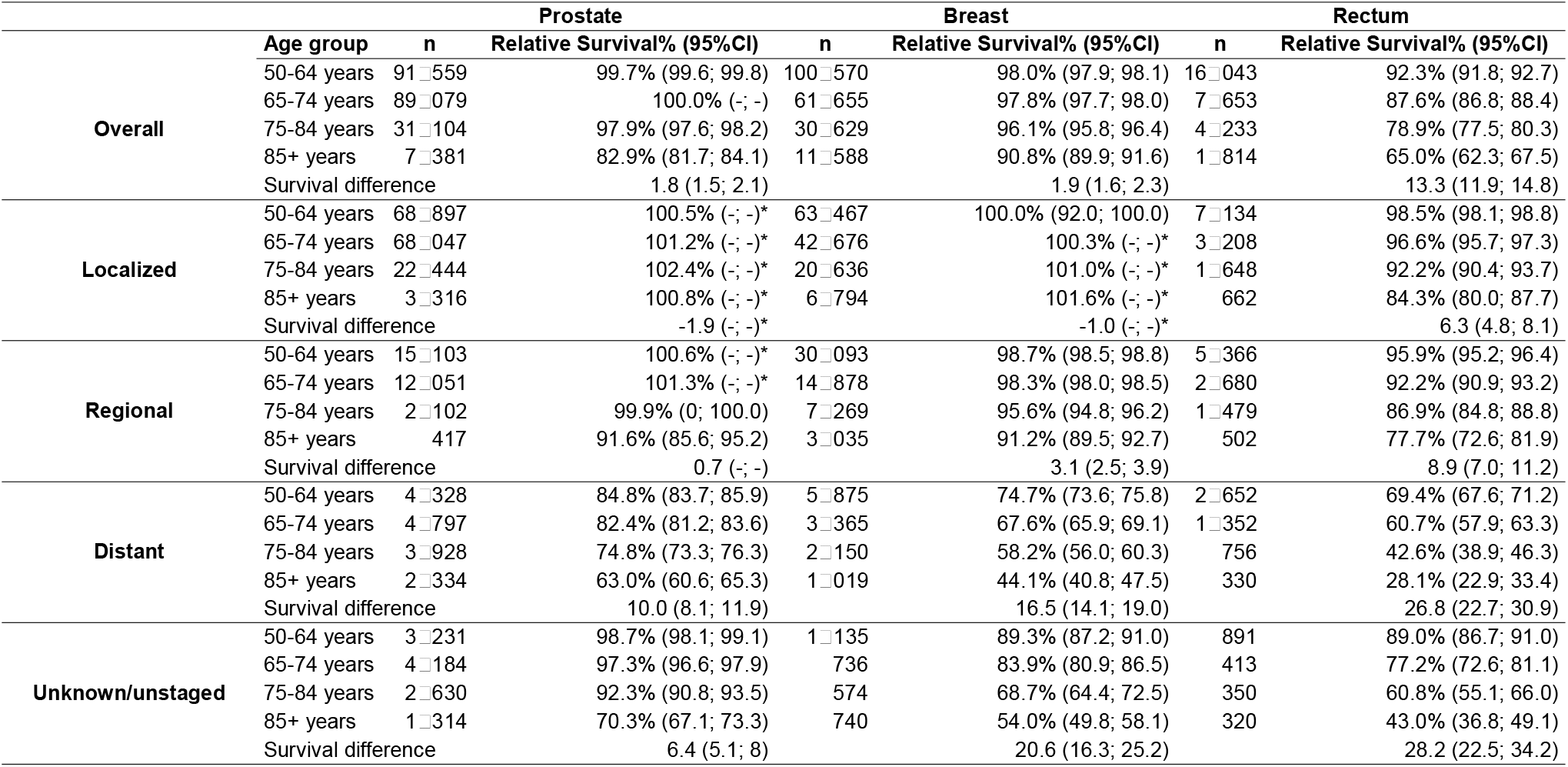
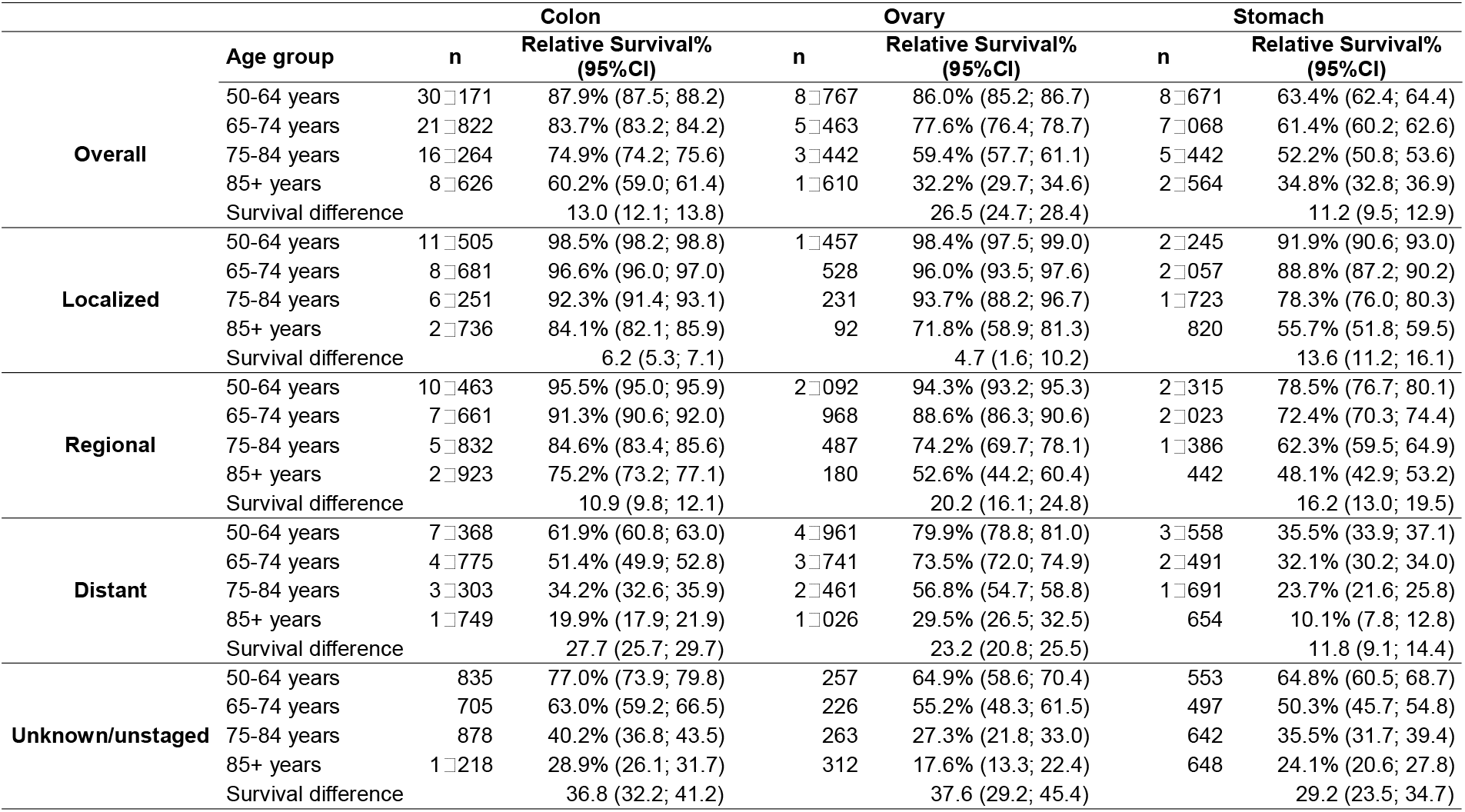
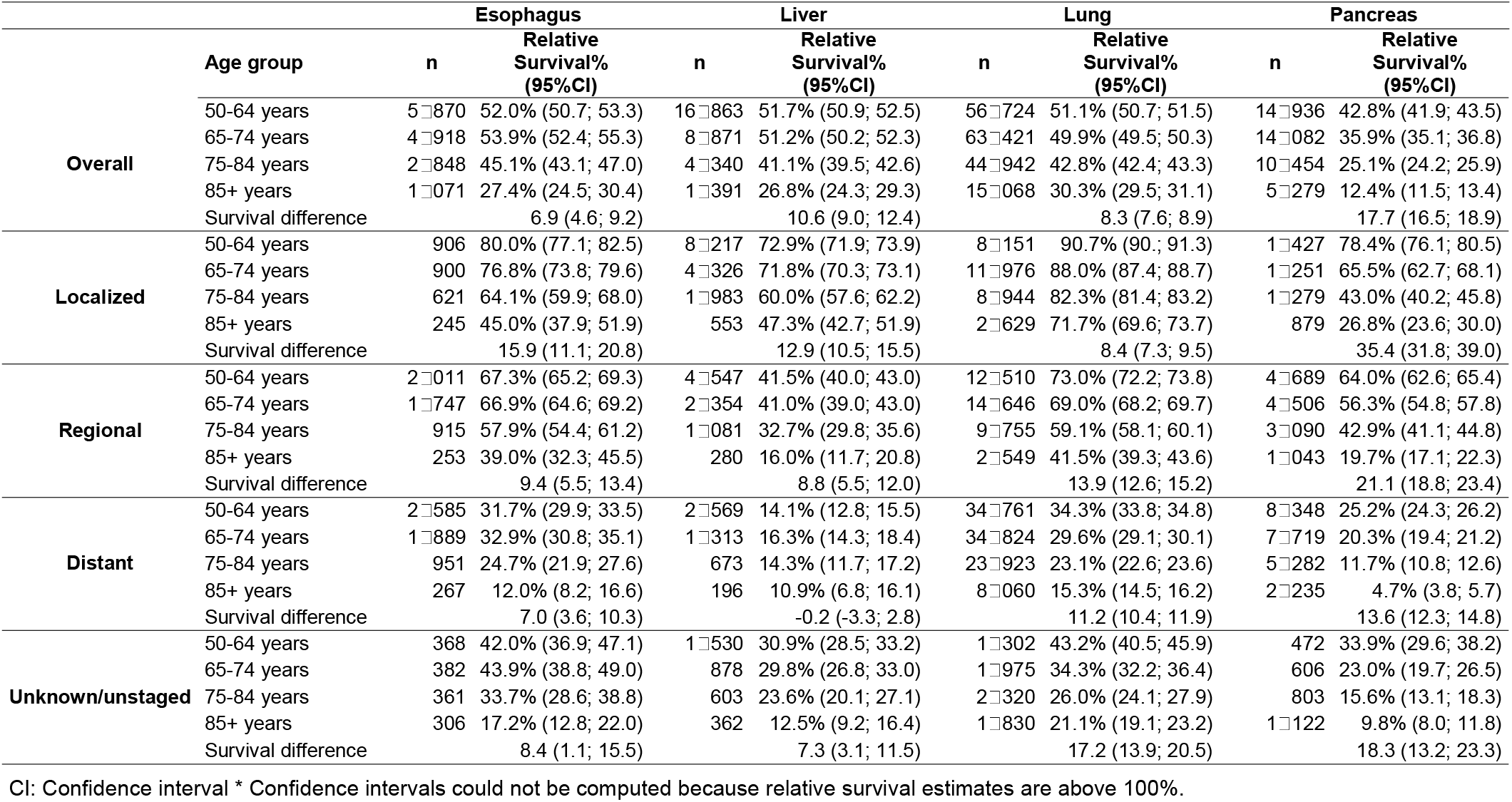
One-year relative survival from cancer by type, stage, and age. Survival differences between those aged 75 to 84 and aged 50 to 64. SEER18 registries, diagnosed 2012 to 2016.

The smallest absolute difference in 1-year RS between the 50 to 64 age group and 75 to 84 age group was observed for prostate and breast cancers with a difference of 1.8 percentage points (95% confidence interval [CI] for prostate: 1.5-2.1), and 1.9 percentage points (95% CI breast: 1.6-2.3), respectively, and the largest was for ovarian cancer with a 27 percentage point difference (95% CI: 25-28, 1-year RS = 86% in the 50 to 64 age group vs. 59% in the 75 to 84 age group). The age-related differences in 1-year RS for other cancers ranged from 7 percentage points for esophageal cancer (95% CI: 5-9) to 18 percentage points for pancreatic cancer (95% CI: 17-19).

### Stage-specific differences (Table 1, Figure 2)

Overall, age-related differences in 1-year RS increased with worsening stage for the better prognosis cancer types (i.e., those with overall 1-year RS exceeding 85%: prostate, breast, colon, rectum, and ovary), while the opposite was observed for all the remaining cancers except stomach cancer. For stomach cancer, age-related differences remained relatively consistent irrespective of stage.

**Figure 2.**
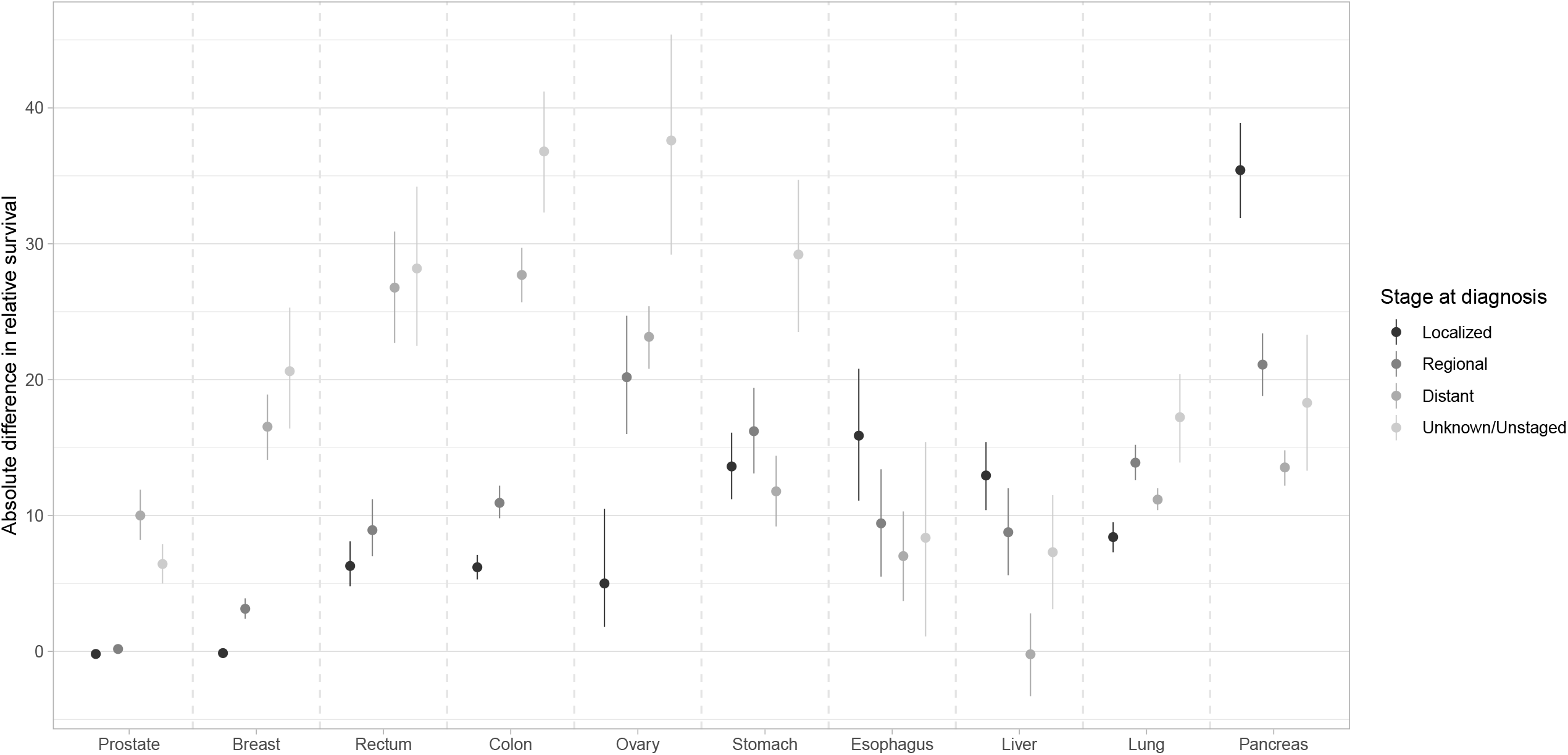
Absolute difference in relative survival between 50 to 64 year olds and 75 to 84 year olds by cancer site and stage. Note: Capped lines are 95% confidence intervals around difference

For localized cancers, the biggest age-related difference in 1-year RS was seen for pancreatic cancer with a difference of over 35 percentage points (95% CI: 32-39) between RS in the 50 to 64 age group (78%) and the 75 to 84 age group (43%). For localized prostate and breast cancers, the age-related difference was negligible.

For regional cancers, age-related differences in 1-year RS were greatest for pancreatic cancer (21 percentage point difference, 95% CI: 19-23) and ovarian cancer (20, 95% CI: 16-25). Differences ranged between 9 and 16 percentage points for stomach, esophagus, liver, and lung cancers.

For distant cancers, the greatest age-related differences in 1-year RS were observed for colon and rectal cancers with differences of 28 (95% CI: 26-30) and 27 (95% CI: 23-31) percentage points respectively, closely followed by ovarian cancer with a difference of 23 points (95% CI: 21-25). For distant liver cancer, there was no difference in survival by age (1-year RS = 14% in 50 to 64 age group and 75 to 84 age group).

For cancers of unknown stage, the age-related difference in 1-year RS ranged between 6 (prostate cancer, 95% CI: 5-8) and 37 percentage points (colon cancer, 95% CI: 32-41).

### Conditional relative survival (Supplemental Table)

Overall, in patients who survived the first year after diagnosis, age-related differences in 1-year relative survival were practically absent for colon, stomach, esophagus, lung cancers and were greatly reduced for ovarian (11%-points vs. 27%-points in the first year), liver (5%-points vs. 11%-points), and pancreatic cancers (6%-points vs. 18%-points).

By stage, age-related differences remained relatively large for distant rectal, colon, and ovarian cancers (9 to 13%-points) as well as for cancers of rectum, colon, ovary, stomach, esophagus, and pancreas that were of unknown stage (10-27%-points).

## Discussion

Using high-quality data from 18 SEER population-based cancer registries, we described overall and stage-specific age-related differences in 1-year RS for 10 cancer types. For some cancers with overall very good prognoses (e.g., localized breast and prostate cancers, 1-year RS >95% for adults aged 50-84) or very poor prognoses (distant stage esophageal and liver cancers, 1-year RS <40%), age-related differences in RS were negligible. Age-related differences were largest for localized pancreatic cancer and unknown stage rectal, colon and ovarian cancers. Age-related differences in survival diminished or were eliminated in the second year after diagnosis conditioning on one-year survival, suggesting that differences arise in the first year after diagnosis. Broad age-related variation in survival by prognosis (i.e., cancer type and/or stage) concentrated in the first year, suggests that the magnitude of age-related survival differences may depend on the extent to which cancers are amenable to treatment.

Our results add to the growing international body of work that describes age-related differences in cancer survival.^6,7,9,22^ Whereas other studies commonly adjust for age at diagnosis, we considered age at diagnosis as an independent axis of investigation. Without measuring age-related differences directly, Zeng et al. reported that improvements in cancer survival between 1990 and 2009 in the US were significantly larger for younger patients with cancer, and that this was particularly the case for colorectal, breast, and early stage liver cancers.^5^ The magnitude of the differences reported here are smaller than those reported for cases diagnosed in France between 1989 and 1997,^6^ but similar to those reported for a pooled European analysis of cases from 2000 to 2002.^7^ The ICBP study compared survival among persons by age splitting at age 75 for all stages combined and, like this study in the US, found large age-related differences in survival for ovarian and pancreatic cancers, but the rank order of cancers by age-related difference varied across countries. Age-related differences in lung cancer survival were high in New Zealand and Norway, for example, and age-related differences in esophageal cancer survival were higher in Ireland and Norway.

With respect to stage, in New Zealand, authors found greater age-related differences in colon cancer survival with worsening stage at diagnosis,^16^ and age-related in differences in lung cancer survival were higher for regional vs. advanced disease,^9^ both patterns being consistent with our results. Given differences in health care systems internationally and potential differences in data quality, collection, and analytical methods, the exact magnitude of age-related differences are difficult to compare.

We observed that when diagnosed early with relatively good prognosis cancers (e.g., localized and regional stage prostate and breast cancers, and localized colorectal and ovarian cancers) or with late-stage poor prognosis cancer (e.g., liver), older adults have similar survival probabilities (when considering differences in life expectancy) as their younger peers. If treatment is a primary driver of age-related difference in cancer survival as suggested by the smaller differences in RS observed in patients surviving the first year, our results would suggest equally tolerated treatment regimens across age groups for the diagnoses with better prognoses. For diagnoses with poorer prognoses, the small differences observed across age groups are likely to be explained by their high lethality regardless of patients’ age.

Age-related differences in RS were greater in good prognosis cancers diagnosed at later stage (e.g., colon or rectal cancers) and poor prognosis cancers diagnosed at early stage (e.g., pancreatic, esophageal, or liver cancers). This observation may be explained by several factors. First, different treatment strategies may be offered or accepted in younger and older patients because of differences in health status or patients’ preferences. Older adults may be less likely to receive curative treatment,^23-29^ and especially in the face of uncertain prognosis, may be more likely to value quality of life over survival.^30^ Second, in the context of a poor evidence base on treatment strategies in older adults,^31^ the effectiveness and toxicity of treatment may differ across age groups. Evidence suggests higher post-treatment mortality and increased all-cause mortality in older patients, particularly in frailer ones.^32^

Factors other than treatment could also contribute to the age-related differences in cancer survival reported here, such as tumor-specific factors. For example, age-related differences in lung cancer survival have been shown to vary by histology, and by subsite in colorectal cancer.^16^ It is also possible that insurance status or more generally access to care may affect age-related differences in survival. Future research should continue to disentangle the role of these factors on age-related differences across a range of cancer types.

As is apparent from Figure 1, age is associated with higher likelihood of having an unstaged cancer.^33^ We observed large age-related differences in cancer survival for colon, rectal, ovarian and stomach cancers of unknown stage. Cancers will have unknown stage if there were insufficient information for the registry to assign stage or may reflect a decision not to pursue a staging workup if the potential harms outweigh the benefits (i.e., due to patient preference, or if the patient is too frail for cancer treatment regardless of stage). In the absence of reasons for missingness and how they vary by age, it is difficult to explain age-related differences in cancer survival for cancer of unknown stage in our study. For example, if all younger patients had missing stage due to data collection errors and all older patients had cancer of unknown stage because it was not investigated (poor health status or patient preferences), the differences in survival between them would be inappropriate to interpret since they represent different types of patients or cancers. While the determinants of unstaged disease have been explored and age has been determined as a primary factor,^34-36^ the determinants of missingness within age categories, and how they differ by age have not been documented and are worthy of future research in order to better interpret results of studies like this one.

This study has limitations, largely as a consequence of its broad scope. Our multi-cancer approach and use of cancer registry data prohibited a detailed analysis of the influence of specific patient, tumor, and/or healthcare system factors that could also explain age-related differences in survival. For example, because of the lack of information about comorbidity, frailty, or other geriatric conditions, it was not possible to distinguish fit versus frail patients. Consequently, we were unable to disentangle which share of the age-related differences in survival observed may be avoidable versus arose as a consequence of differences in baseline health status. Despite these limitations, the paper provides insight on the role of stage in age-related disparities and how this varies by cancer type, knowledge which can be mobilized to priority efforts to narrow age gaps in survival.

With respect to methods, the estimation of relative survival relies on background mortality rates obtained from lifetables and implies that in the absence of cancer, people in our study cohort would have the same mortality risk as the general population. While the US life tables account for differences in age, sex, ethnicity and county SES between cancer patients and the general population, they are not specific to other factors influencing prognosis and mortality (such as comorbidities and smoking), for which the prevalence may differ between cancer patients and the general population. Consequently, age-related differences in relative survival estimates may be underestimated.^37^ In a recent study describing age-related differences in lung cancer survival, correction of life tables for smoking did change net survival estimates, but did not meaningfully change age-related differences in survival,^9^ our main outcome of interest in the present study.

In this population-based study, we confirmed that cancer survival in older adults tends to be poorer even after accounting for differences in background mortality for a range of common cancers and we illustrated that the patterns of age-related differences varied by cancer type and stage and aligning with prognosis. More detailed studies of the determinants of age-related differences in cancer survival, considering characteristics of the patients, their cancers, and their treatment, are necessary to achieve better outcomes in older adults.

## Data Availability

Data and relative survival estimates were retrieved through SEER*Stat software version 8.3.9 (National Cancer Institute).

## Abbreviations

CI: Confidence Interval
ICBP: International Cancer Benchmarking Partnership
RS: Relative Survival
SEER: Surveillance Epidemiology and End Results
SES: socioeconomic status

## Acknowledgment

Authors thank Dr Hadrien Charvat for his guidance regarding calculating 95% confidence interval around differences in relative survival.

## Author contributions

The work reported in the paper has been performed by the authors, unless clearly specified in the text.

### CRediT author statement

Diana Withrow and Sophie Pilleron: conceptualization, methodology, formal analysis, writing – original draft, project administration; Melisa Wong, Eva Morris and Brian Nicholson: interpretation, writing – review and editing.

## Data accessibility

Data that support this study are available via the SEER*Stat page of the NIH website: https://seer.cancer.gov/seerstat/.

