## Supplemental Table for "Age-related differences in cancer relative survival in the US: a SEER-18 analysis"

**Supplemental Table 1.** One-year relative survival from cancer conditioning on surviving the first year of diagnosis by type, stage, and age. Survival differences between those aged 75 to 84 and aged 50 to 64. SEER18 registries, diagnosed 2012 to 2016

**Supplemental Table 1.** One-year relative survival from cancer conditioning on surviving the first year of diagnosis by type, stage, and age. Survival differences between those aged 75 to 84 and aged 50 to 64. SEER18 registries, diagnosed 2012 to 2016. Continued on next page.

|  |  | **Prostate** | | | **Breast** | | | **Rectum** | | | **Colon** | | | **Ovary** | |
| --- | --- | --- | --- | --- | --- | --- | --- | --- | --- | --- | --- | --- | --- | --- | --- |
|  | **Age group** | **n** | **RS (95%CI)** | **n** | | **RS (95%CI)** | **n** | | **RS (95%CI)** | **n** | | **RS (95%CI)** | **n** | | **RS (95%CI)** |
| **Overall** | 50-64 years | 87601 | 99.4 (99.3; 99.5) | 94712 | | 97.9 (97.8; 98.0) | 14249 | | 93.2 (92.7; 93.6) | 25432 | | 91.9 (91.5; 92.3) | 7285 | | 88.7 (87.9; 89.5) |
|  | 65-74 years | 85173 | 99.8 (99.6; 99.8) | 58061 | | 98.6 (98.5; 98.8) | 6456 | | 91.5 (90.7; 92.3) | 17552 | | 91.9 (91.4; 92.3) | 4101 | | 84.6 (83.4; 85.8) |
|  | 75-84 years | 28243 | 98.2 (97.9; 98.6) | 27720 | | 97.8 (97.5; 98.1) | 3128 | | 89.2 (87.7; 90.5) | 11355 | | 91 (90.3; 91.7) | 1945 | | 77.6 (75.4; 79.7) |
|  | 85+ years | 5185 | 90.3 (88.7; 91.6) | 9145 | | 95.2 (94.2; 96.1) | 1017 | | 78.7 (75.0; 81.9) | 4495 | | 89.8 (88.3; 91.2) | 458 | | 63.3 (57.6; 68.5) |
|  | Difference |  | 1.2 (0.9; 1.6) |  | | 0.1 (-0.2; 0.4) |  | | 4 (2.6; 5.6) |  | | 0.9 (0.1; 1.7) |  | | 11.1 (8.9; 13.5) |
| **In situ/localized** | 50-64 years | 66415 | 100 (-; -)* | 60874 | | 99.8 (99.7; 99.8) | 6723 | | 98.8 (98.4; 99.1) | 10814 | | 99.1 (98.8; 99.3) | 1371 | | 98.5 (97.4; 99.1) |
|  | 65-74 years | 65858 | 100.1 (-; -)* | 41203 | | 100 (-; -) | 2977 | | 98.1 (97.2; 98.7) | 8052 | | 98.5 (98.0; 98.9) | 489 | | 98.6 (95.9; 99.5) |
|  | 75-84 years | 21335 | 100.2 (-; -)* | 19657 | | 100.1 (-; -) | 1427 | | 96.5 (94.5; 97.8) | 5380 | | 98 (97.1; 98.6) | 198 | | 99.4 (87.7; 100) |
|  | 85+ years | 2834 | 100.3 (-; -)* | 6010 | | 100.1 (-; -) | 484 | | 89.4 (84.0; 93.1) | 1980 | | 98.7 (95.3; 99.7) | 56 | | 91.9 (77.4; 97.2) |
|  | Difference |  | - |  | | - |  | | 2.3 (1.0; 4.4) |  | | 1.1 (0.4; 2.0) |  | | -0.9 (-2.2; 10.6) |
| **Regional** | 50-64 years | 14615 | 99.9 (99.4; 100) | 28646 | | 97.0 (96.7; 97.2) | 4984 | | 94.7 (93.9; 95.4) | 9567 | | 95.1 (94.6; 95.6) | 1897 | | 95.5 (94.3; 96.4) |
|  | 65-74 years | 11681 | 100.1 (-; -) | 14076 | | 97.5 (97.1; 97.8) | 2380 | | 92.1 (90.7; 93.3) | 6712 | | 94.3 (93.5; 94.9) | 825 | | 90.5 (88.0; 92.6) |
|  | 75-84 years | 1949 | 98.5 (96.5; 99.4) | 6516 | | 94.4 (93.6; 95.2) | 1206 | | 91.2 (88.7; 93.1) | 4586 | | 91.4 (90.2; 92.4) | 344 | | 93.5 (88.8; 96.3) |
|  | 85+ years | 325 | 90.1 (82.7; 94.4) | 2399 | | 89.4 (87.2; 91.2) | 338 | | 80.5 (73.7; 85.7) | 1913 | | 91.6 (89.1; 93.5) | 83 | | 81.3 (66.7; 89.9) |
|  | Difference |  | 1.4 (0.4; 3.3) |  | | 2.5 (1.8; 3.4) |  | | 3.5 (1.5; 6.0) |  | | 3.7 (2.6; 5.0) |  | | 2.0 (-1.0; 6.8) |
| **Distant** | 50-64 years | 3533 | 79.6 (78.1; 81.0) | 4264 | | 79.0 (77.6; 80.2) | 1792 | | 67.1 (64.6; 69.3) | 4444 | | 67.9 (66.4; 69.3) | 3857 | | 82.5 (81.1; 83.7) |
|  | 65-74 years | 3756 | 79.1 (77.5; 80.5) | 2203 | | 79.2 (77.3; 81.0) | 797 | | 67.2 (63.4; 70.6) | 2366 | | 64.8 (62.7; 66.9) | 2668 | | 80.6 (78.9; 82.2) |
|  | 75-84 years | 2716 | 74.2 (72.1; 76.1) | 1180 | | 74.7 (71.7; 77.4) | 297 | | 53.9 (47.4; 59.9) | 1061 | | 58.5 (55.1; 61.7) | 1336 | | 71.8 (68.9; 74.4) |
|  | 85+ years | 1246 | 70.4 (66.8; 73.6) | 395 | | 65.7 (59.5; 71.3) | 80 | | 49 (35.7; 61.1) | 304 | | 48.7 (41.7; 55.4) | 271 | | 54.8 (47.5; 61.6) |
|  | Difference |  | 5.4 (3.0; 7.9) |  | | 4.2 (1.2; 7.5) |  | | 13.2 (6.7; 20.1) |  | | 9.4 (5.9; 13.1) |  | | 10.7 (7.8; 13.8) |
| **Unknown/unstaged** | 50-64 years | 3038 | 98.4 (97.6; 98.9) | 928 | | 92.1 (90.0; 93.9) | 750 | | 95.1 (93.0; 96.6) | 607 | | 87.5 (84.3; 90.1) | 160 | | 81.5 (73.9; 87.1) |
|  | 65-74 years | 3878 | 97.9 (97.1; 98.6) | 579 | | 92.3 (89.3; 94.4) | 302 | | 86.7 (81.5; 90.5) | 422 | | 80.4 (75.7; 84.3) | 119 | | 79.0 (69.5; 85.8) |
|  | 75-84 years | 2243 | 94.9 (93.2; 96.2) | 367 | | 86.4 (81.3; 90.1) | 198 | | 77.7 (69.9; 83.7) | 328 | | 75.2 (69.2; 80.2) | 67 | | 54.4 (40.5; 66.4) |
|  | 85+ years | 780 | 84.9 (80.4; 88.4) | 341 | | 80.6 (73.7; 85.8) | 115 | | 49.1 (37.4; 59.7) | 298 | | 56.9 (49.5; 63.6) | 48 | | 42.0 (25.4; 57.8) |
|  | Difference |  | 3.5 (2.0; 5.2) |  | | 5.8 (1.5; 11.1) |  | | 17.4 (11.1; 25.3) |  | | 12.3 (6.4; 18.9) |  | | 27.1 (13.1; 42.1) |

|  |  | **Stomach** | | **Esophagus** | | **Liver** | | **Lung** | | **Pancreas** | |
| --- | --- | --- | --- | --- | --- | --- | --- | --- | --- | --- | --- |
|  | **Age group** | **n** | **RS (95%CI)** | **n** | **RS (95%CI)** | **n** | **RS (95%CI)** | **n** | **RS (95%CI)** | **n** | **RS (95%CI)** |
| **Overall** | 50-64 years | 5251 | 74.1 (72.8; 75.4) | 2981 | 66.0 (64.2; 67.8) | 8465 | 72.9 (71.9; 73.9) | 28207 | 70.6 (70.1; 71.2) | 6199 | 57.4 (56.1; 58.7) |
|  | 65-74 years | 4140 | 77.9 (76.4; 79.2) | 2560 | 68.4 (66.4; 70.4) | 4362 | 72.4 (70.9; 73.9) | 30625 | 72.5 (71.9; 73.0) | 4883 | 54.1 (52.6; 55.6) |
|  | 75-84 years | 2640 | 76.0 (74.0; 77.8) | 1204 | 64.5 (61.3; 67.5) | 1659 | 67.9 (65.3; 70.4) | 18155 | 70.8 (70.0; 71.5) | 2470 | 51.6 (49.3; 53.8) |
|  | 85+ years | 779 | 67.0 (62.7; 71) | 255 | 51.8 (44.2; 58.9) | 328 | 57.2 (50.4; 63.4) | 4000 | 66.3 (64.4; 68.1) | 572 | 40.3 (35.5; 45.1) |
|  | Difference |  | -1.9 (-4.1; 0.4) |  | 1.5 (-2.0; 5.2) |  | 5.0 (2.3; 7.8) |  | -0.1 (-1.1; 0.8) |  | 5.8 (3.3; 8.4) |
| **In situ/localized** | 50-64 years | 1968 | 93.0 (91.6; 94.2) | 706 | 85.1 (82.0; 87.7) | 5833 | 79.3 (78.1; 80.4) | 7134 | 91.3 (90.5; 92.0) | 1076 | 84.8 (82.3; 87.0) |
|  | 65-74 years | 1740 | 93.1 (91.5; 94.4) | 669 | 87.1 (83.8; 89.7) | 2979 | 78.4 (76.7; 80.0) | 10200 | 90.2 (89.5; 90.9) | 788 | 82.5 (79.3; 85.2) |
|  | 75-84 years | 1254 | 90.1 (87.8; 92.1) | 372 | 85.7 (80.6; 89.6) | 1109 | 74.0 (70.8; 76.8) | 6921 | 85.9 (84.9; 86.9) | 516 | 69.9 (65.1; 74.2) |
|  | 85+ years | 401 | 79.5 (73.6; 84.2) | 95 | 67.5 (53.8; 78.0) | 229 | 63.7 (55.5; 70.8) | 1652 | 82.6 (79.8; 85.0) | 205 | 48.7 (40.0; 56.9) |
|  | Difference |  | 2.9 (0.5; 5.5) |  | -0.6 (-5.6; 5.1) |  | 5.3 (2.2; 8.6) |  | 5.4 (4.1; 6.6) |  | 14.9 (10.0; 20.2) |
| **Regional** | 50-64 years | 1746 | 70.9 (68.5; 73.1) | 1324 | 69.1 (66.3; 71.7) | 1827 | 60.9 (58.5; 63.2) | 8911 | 76.9 (75.9; 77.8) | 2919 | 58.5 (56.6; 60.4) |
|  | 65-74 years | 1404 | 74.9 (72.3; 77.2) | 1130 | 67.9 (64.8; 70.9) | 928 | 63.5 (60.0; 66.8) | 9794 | 76.2 (75.2; 77.1) | 2447 | 54.4 (52.2; 56.5) |
|  | 75-84 years | 803 | 69.7 (65.9; 73.2) | 498 | 59.2 (54.0; 64.0) | 331 | 60.6 (54.5; 66.2) | 5443 | 72.6 (71.2; 73.9) | 1257 | 49.7 (46.6; 52.8) |
|  | 85+ years | 186 | 57.8 (48.5; 66) | 87 | 47.9 (35.1; 59.5) | 40 | 34.9 (17.8; 52.6) | 930 | 64.8 (60.9; 68.5) | 180 | 36.3 (27.9; 44.8) |
|  | Difference |  | 1.2 (-3.0; 5.6) |  | 9.9 (4.4; 15.7) |  | 0.3 (-5.7; 6.8) |  | 4.3 (2.6; 6.0) |  | 8.8 (5.2; 12.5) |
| **Distant** | 50-64 years | 1210 | 47.2 (44.2; 50.2) | 804 | 44 (40.3; 47.6) | 350 | 42.8 (37.1; 48.3) | 11629 | 53.5 (52.5; 54.4) | 2051 | 41.1 (38.8; 43.3) |
|  | 65-74 years | 761 | 47.4 (43.5; 51.2) | 605 | 51.2 (46.8; 55.4) | 204 | 44.7 (37.1; 52.0) | 9983 | 52.0 (51.0; 53.1) | 1514 | 38.4 (35.7; 41.1) |
|  | 75-84 years | 371 | 47.6 (41.9; 53.0) | 220 | 47.9 (40.4; 55.0) | 88 | 39.8 (28.9; 50.5) | 5224 | 50.4 (48.8; 51.9) | 581 | 38.1 (33.7; 42.5) |
|  | 85+ years | 58 | 40.0 (25.3; 54.2) | 28 | 47.6 (25.8; 66.6) | 19 | 32.8 (9.9; 58.4) | 1084 | 47.5 (43.9; 50.9) | 93 | 34.1 (23.5; 44.9) |
|  | Difference |  | -0.3 (-6.5; 6.1) |  | -3.9 (-11.8; 4.4) |  | 3 (-9.0; 15.2) |  | 3.1 (1.3; 4.9) |  | 3.0 (-1.9; 7.9) |
| **Unknown/unstaged** | 50-64 years | 327 | 77.4 (72.1; 81.8) | 147 | 69.0 (60.1; 76.3) | 455 | 62.9 (57.9; 67.4) | 533 | 66.6 (62.1; 70.7) | 153 | 66.6 (57.9; 73.9) |
|  | 65-74 years | 235 | 81.0 (74.7; 85.9) | 156 | 60.1 (51.1; 67.9) | 251 | 57.5 (50.4; 63.9) | 648 | 59.1 (54.9; 63.1) | 134 | 57.2 (47.5; 65.7) |
|  | 75-84 years | 212 | 65.5 (57.7; 72.3) | 114 | 49.2 (38.5; 59.0) | 131 | 56.3 (46.7; 64.9) | 567 | 58.4 (53.7; 62.7) | 116 | 56.5 (45.7; 66.0) |
|  | 85+ years | 134 | 51.7 (41.0; 61.4) | 45 | 29.2 (15.6; 44.3) | 40 | 51.6 (32.6; 67.6) | 334 | 54.2 (47.6; 60.4) | 94 | 36.1 (24.9; 47.4) |
|  | Difference |  | 11.9 (3.3; 20.9) |  | 19.9 (6.7; 32.8) |  | 6.5 (-3.3; 17.2) |  | 8.3 (2.0; 14.5) |  | 10.1 (-2.6; 23.1) |

CI: Confidence interval * Confidence intervals could not be computed because relative survival estimates are above 100%.
